# Relationship of preoperative oral hypofunction with prognostic nutritional index in gastric cancer

**DOI:** 10.1101/2022.08.26.22279280

**Authors:** Atsushi Abe, Atsushi Nakayama, Yuya Otsuka, Kanae Shibata, Yoshihito Matsui, Yu Ito, Hiroki Hayashi, Moeko Momokita, Shinichi Taniguchi

## Abstract

We recruited patients with perioperative gastric cancer and examined preoperative oral hypofunction and its relationship with prognostic nutritional index. This cross-sectional study analysed 95 patients who underwent oral function management. We assessed the following parameters: body mass index, stage of gastric cancer, C-reactive protein, total lymphocyte count, albumin, and prognostic nutritional index. The patients were divided into two groups: prognostic nutritional index <45 and prognostic nutritional index >45. Logistic regression analysis was used to assess the association between the measurements of oral function and the prognostic nutritional index.A univariate analysis of factors associated with decreased oral function and prognostic nutritional index showed significant differences in C-reactive protein, neutrophils and tongue pressure (p<0.01) between the two groups. However, oral hygiene, oral dryness, occlusal force, tongue-lip motor function, masticatory function, and swallowing function were not significantly different. Multivariate analysis showed that C-reactive protein (odds ratio: 0.12, 95% confidence interval: 0.30–0.45, p<0.01) and tongue pressure (odds ratio: 3.62, 95% confidence interval: 1.04–12.60, p<0.05) were independent risk factors for oral hypofunction. Oral function is decreased in perioperative patients with gastric cancer, and decreased tongue pressure is associated with decreased prognostic nutritional index.

## Introduction

Nutritional status and systemic inflammatory response in patients with gastric cancer affect disease recovery, reduce vand cause postoperative complications and hospital mortality [1–3]. Systemic inflammatory responses and malnutrition are caused by inadequate dietary intake, reduced activity, oxidative stress, increased inflammatory cytokines, and hormonal imbalances [4,5]. The indicators calculated from the preoperative blood sampling data are used as an objective method of assessing the nutritional status [6]. Currently, the prognostic nutritional index (PNI) is considered to be associated with prognosis in several cancers [3,7–10]. The PNI is calculated from serum albumin levels and total lymphocyte counts in peripheral blood, and has been proposed for assessing the perioperative immunonutritional status and surgical risk of patients scheduled for gastrointestinal surgery. Low PNI may lead to increased postoperative complications and worsened prognosis [11].

Oral function plays a major role in the feeding and swallowing process, and oral dysfunction worsens the nutritional status [12–14] and is significantly associated with sarcopenia, low nutrition, and hypotrophy. Decreased oral function decreases the quantity and quality of food intake, leading to muscle hypotrophy and weight loss, which lead to a negative cascade of events that promote further progression of hypotrophy and sarcopenia. The individual functions of the parts of the oral cavity work complementarily; therefore, the decline in oral function is not detected early and progresses gradually. Therefore, oral hypofunction (OHF) assessment has been proposed to quantitatively evaluate functional decline, which will lead to early detection of oral hypofunction [15,16]. The diagnosis of oral hypofunction is based on seven tests, and a diagnosis is made when the results of three or more of the seven tests are below normal values. The tests include the assessment of oral moisture, bite force, tongue-lip motor function, tongue pressure, masticatory function, and swallowing function. Previous studies have associated poor oral hypofunction with malnutrition, sarcopenia, and frailty [17–20]. However, the studies were conducted on institutionalised elderly patients, and few studies have correlated preoperative nutritional status and oral function decline in patients with gastric cancer. Therefore, we investigated the relationship between the changes in preoperative oral function and the PNI, an immunonutritional index, in patients with gastric cancer, since a decline in preoperative oral function may contribute to postoperative complications. Furthermore, we investigated the correlation between PNI and the seven factors related to oral hypofunction.

## Materials and Methods

### Participants

A cross-sectional analysis of 112 patients diagnosed with primary gastric cancer who underwent oral functional management at the Nagoya Ekisaikai Hospital from 1 January 2014 to 31 December 2021 was conducted. The inclusion criteria were as follows: (1) no history of other cancers, (2) histopathologically diagnosed gastric cancer, (3) no history of radiation therapy and/or chemotherapy, and (4) available preoperative blood test results. The exclusion criteria included the following: (1) history of oral and pharyngeal cancer and aborted surgery, (2) patients with gastrointestinal stromal tumours, (3) recurrence, (4) metabolic diseases (e.g. diabetes), and (5) Sjogren’s syndrome; and (6)17 patients with missing data. Thus, 95 patients (38 males and 57 females) were enrolled in the study. Patients’ clinical data were recorded using a chart review. No a priori sample size calculation was required due to the retrospective observational study design. This research was independently reviewed and approved by the Ethics Committee of the Nagoya Ekisaikai Hospital (approval number 2021-048). The study was performed in accordance with the Strengthening the Reporting of Observational Studies in Epidemiology (STROBE) guidelines and the principles of the Declaration of Helsinki. The STROBE checklist is shown in Fig 1. The experiments were undertaken with the understanding and written consent of each subject and according to the above mentioned principles.

**Fig 1.**
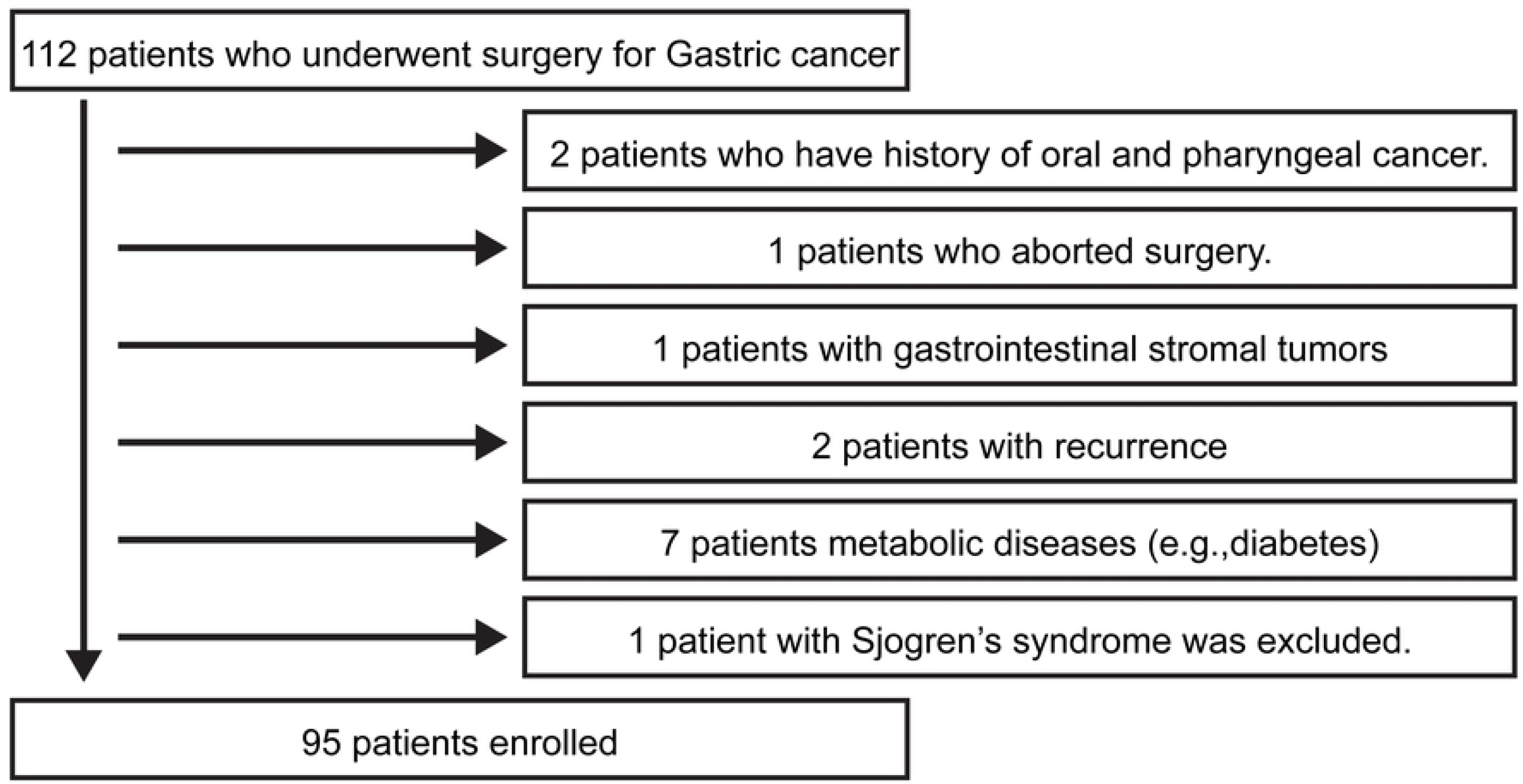
Flow diagram of exam selection.

### Evaluation and general status of patients with gastric cancer

Patients with gastric cancer were diagnosed by endoscopy, computed tomography, and magnetic resonance imaging in the gastrointestinal surgery department of Nagoya Ekisaikai Hospital. Nutritional status was determined by calculating the body mass index (BMI), and albumin (ALB), C-reactive protein (CRP), and total lymphocyte counts were measured by preoperative blood tests. All blood specimens were collected at least one month prior to the preoperative examination for gastric cancer surgery. The patients’ height and weight were measured during their first visit to our clinic. The PNI indicates the nutritional status and systemic inflammatory response. The PNI is a simple scoring system and is calculated based on the serum ALB level and peripheral blood lymphocyte count [21] by the following formula: PNI = [10×serum albumin level (g/dL)] + 0.005×total peripheral lymphocyte count (per mm^3^)].

The PNI is typically quantitative and estimates postoperative surgical complications in surgical patients, with a PNI >45 indicating surgery, 45–40 indicating caution during surgery, and <40 indicating a contraindication for surgery [22,23].

### Evaluation of oral condition

Oral examinations were performed by two calibrated dentists with more than seven years of clinical experience. Based on the diagnostic criteria of the Japanese Society of Geriatric Dentistry, seven sub-symptoms of OHF were measured and evaluated at the dental outpatient clinic on the day or two days before the surgery. The seven items include: (1) oral hygiene, (2) oral dryness, (3) occlusal force, (4) tongue-lip motor function, (5) tongue pressure, (6) masticatory function, and (7) swallowing function.

To assess oral hygiene, the degree of tongue coating was visually evaluated by the tongue coating index (TCI) [24]. The tongue surface was divided into nine sections, and the degree of tongue coating in each section was rated on a scale of 0 - 2, with 0 indicating the least amount of tongue coating. The total score for all the sections was calculated as the TCI. A total score of 9 or more was considered poor oral hygiene [16]. The percentage TCI was calculated as the sum of the scores divided by the maximum score multiplied by 100%. Oral dryness was measured by the wetness of the oral mucosa using an oral moisture meter (Mucus®, Life Co., Ltd., Saitama, Japan) [25,26]. A moisture meter reading of <27 was considered as oral dryness. A tongue pressure probe attached to a digital tongue pressure measuring device (JMS Tongue Depressor®, G.C., Tokyo, Japan, JMS) was pressed against the tongue and palate with maximum force. A mean value of <30 kPa was defined as low tongue pressure [14,27]. The participants were instructed to clench the pressure-sensitive film with their teeth (Dental Prescale II®, G.C. Co., Ltd., Tokyo, Japan) for 3 s, and the colour change on the pressure-sensitive film was analysed and converted into occlusal force using the supplied software. A bite force measurement of <200 N was considered as a decreased bite force [16]. The tongue and lip movement function was measured by instructing the participants to pronounce the sounds /pa/, /ta/, and /ka/ as many times as possible in 5 s, and the number of consecutive pronunciations of /pa/, /ta/, and /ka/ was measured with an oral function measurement device (Kenkuchi-kun Handy, Takei Kikai Kogyo, Niigata, Japan). For the masticatory function, the participants chewed 2 g of gummy jelly (Glucoram, G.C., Tokyo, Japan) for 20 s, and the gummy jelly was gargled with 10 mL of water. The chewed gummies and water were drained through a filtration mesh, and the solution that passed through the mesh was sampled with an applicator to measure the amount of glucose that eluted with a chewing ability testing device (Glucosensor GS-II, G.C., Tokyo, Japan). A glucose density of <100 mg/dL was considered as masticatory dysfunction [28]. Swallowing function was evaluated using the Swallowing Screening Questionnaire (a 10-item Eating Assessment Tool [EAT-10], 40-point scale); a total score of 3 or more was considered as a decline in swallowing function [16].

### Data analysis

The normality of the PNI and patient characteristic data were assessed using the Shapiro–Wilk test. Fisher’s exact test was used for normally distributed data, while Mann–Whitney U test was used for non-normally distributed data. A PNI of 40 or less was used as the objective variable, and five patient characteristics variables (age, gender, smoking, alcohol), the number of current teeth, Eichner classification, and the seven variables that constitute the oral function test were used as explanatory variables in a logistic regression analysis using the stepwise variable reduction method (Wald; probability of elimination: 0.10). The Wald test was applied to test the regression coefficients. The significance level for all statistical analyses was set at 5% on both sides, and the analyses were conducted by forcibly selecting explanatory variables. All statistical analyses were performed using EZR (Saitama Medical Center, Jichi Medical University, Japan) [29].

## Results

### Clinical characteristics of the patients

Among the 112 patients with gastric cancer who consented to the study, 95 (38 males and 57 females) were included in the present analysis. The characteristics of the patients are shown in Table 1. The age of the participants ranged from 37 - 89 years with a mean age ± standard deviation (SD) of 67.2 ± 13.2 years; their BMI ranged from 14.8 - 43.2 kg/m^2^ with a mean ± SD of 22.4 ± 4.1 kg/m^2^. The number of patients with stage I, II, III, and IV gastric cancer was 33 (34.7%), 25 (26.3%), 21 (22.1%), and 16 (16.8%), respectively. No significant between-group differences were found in gender, age, stage of gastric cancer, or BMI. Patients with PNI <45 (24 patients, 25.3%) were classified into the low PNI group and those with PNI greater than 45 (71 patients, 74.7%) into the high PNI group [30].

**Table 1.**
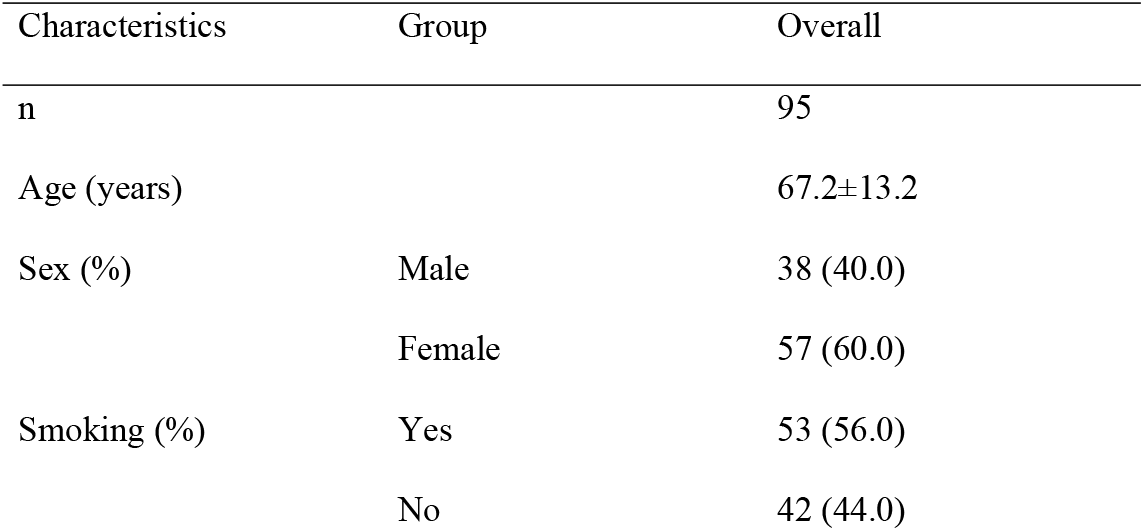
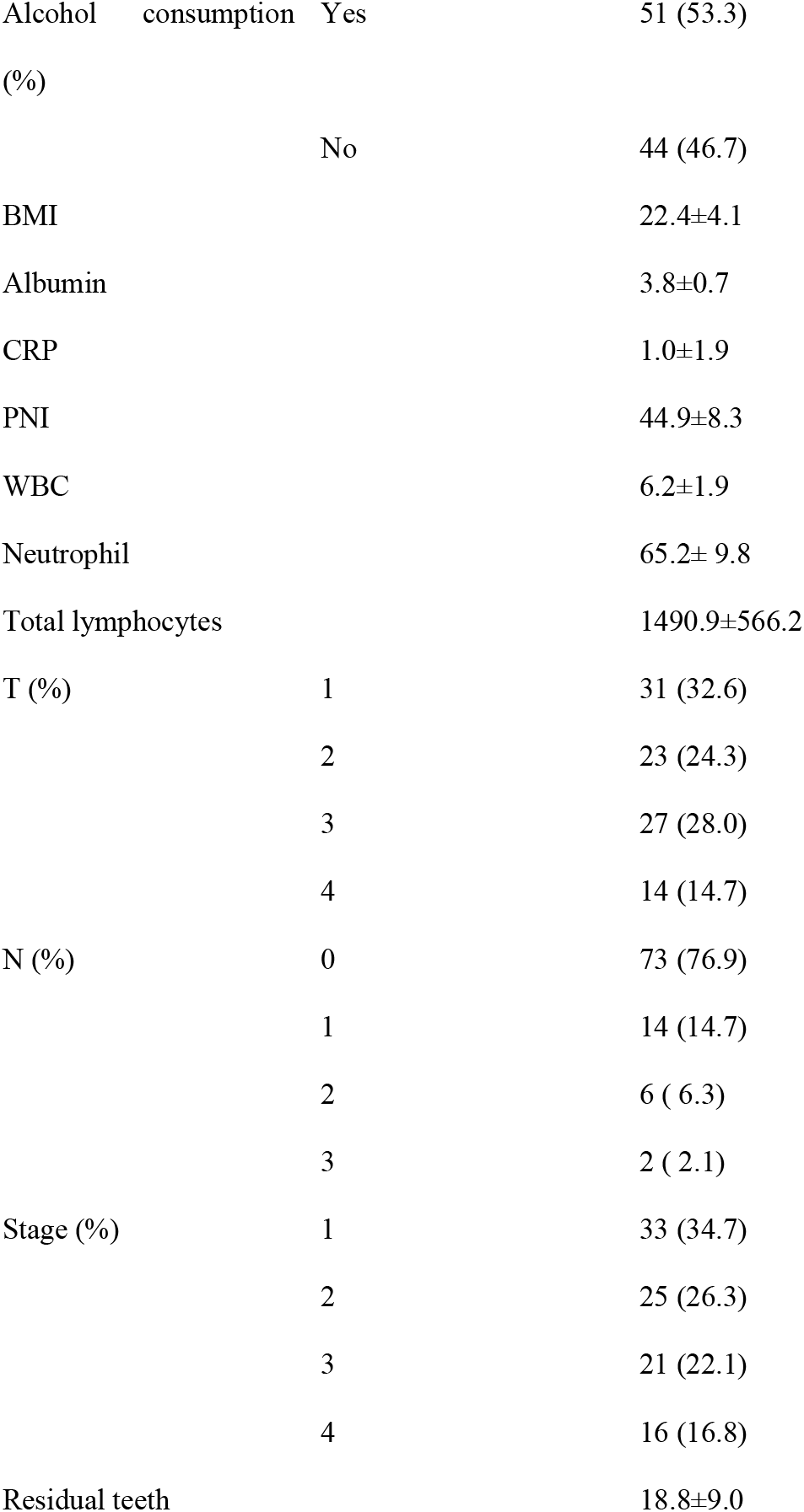
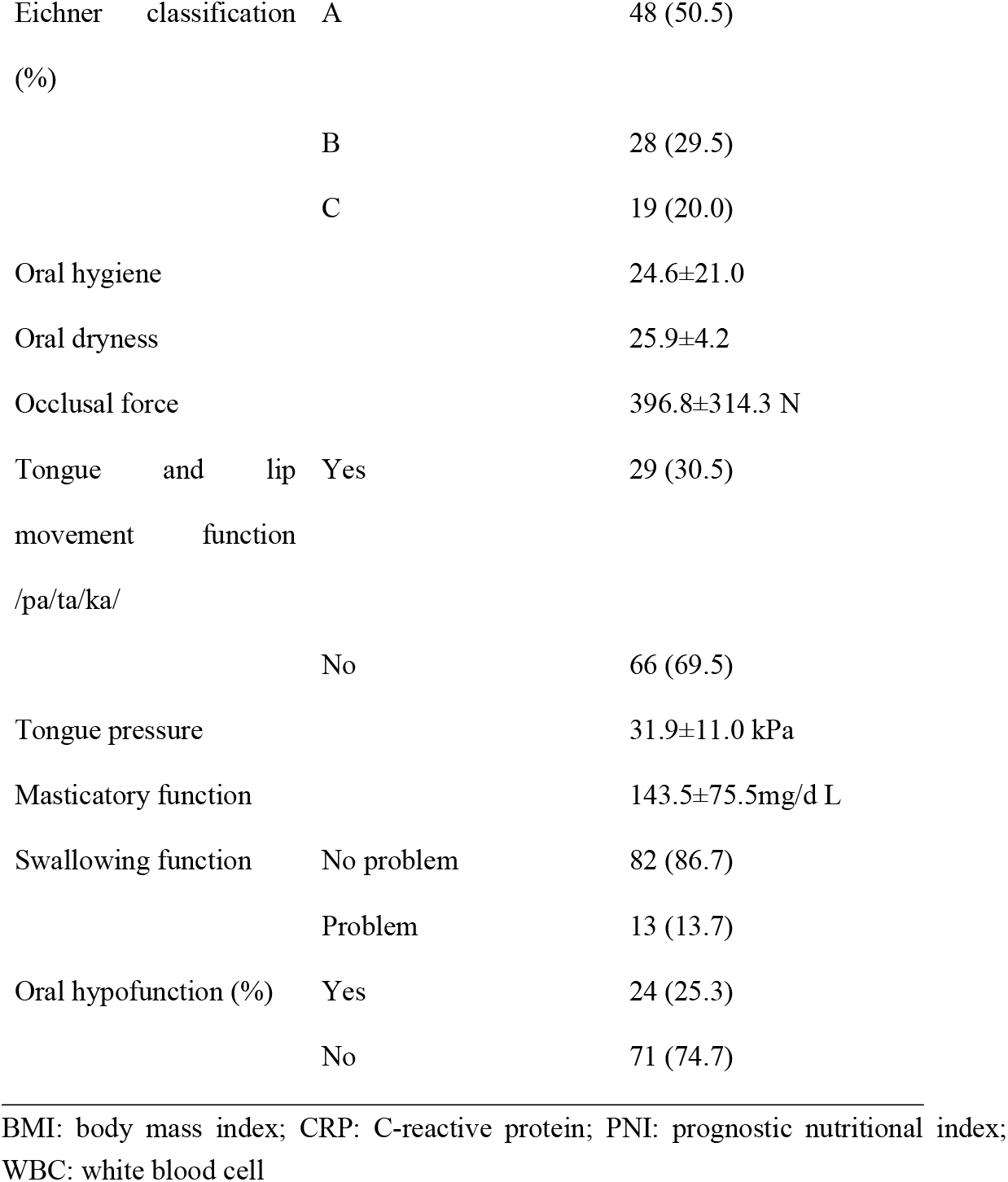
Patient background.

### Oral function

The percentage TCI ranged from 0 - 100% (24.6±21.0%), oral dryness ranged from 8.2 - 32.7 (25.9± 4.2), tongue pressure ranged from 6 - 62.1 kPa (31.9 ± 11.0 kPa), and the occlusal force ranged from 44.9 - 1585.6 N (396.8 ± 314.3 N). The mean value of tongue and lip movement function was less than 6 times in 66 patients (69.5%). Decreased masticatory function ranged from 24.0 to 494.0 mg/dL (143.5± 75.5 mg/dL). The total score for decreased swallowing function was 0 - 14, with 82 patients (86.3%) having a score of 3 or less and 13 patients (13.7%) having a score of >3 In addition, 24 patients (25.3%) had dysfunction in more than three items of oral function and were considered to have OHF. We performed univariate analysis of factors related to PNI for oral dysfunction. The following variables were analysed: age, gender, stage of gastric cancer, CRP, oral hygiene, oral dryness, tongue pressure, occlusal force, tongue and lip movement function, masticatory function, and swallowing function. High oral contamination, such as a TCI of >50%, was observed in four patients in the low PNI group and 12 patients in the high PNI group. Dry mouth was observed in 10 patients in the low PNI group and 23 patients in the high PNI group. Low tongue pressure was observed in 15 patients in the low PNI group and 24 patients in the high PNI group. Decreased occlusal force was observed in 11 patients in the low PNI group and 19 patients in the high PNI group. Decreased tongue and lip motor function was found in 18 patients in the low PNI group and 48 patients in the high PNI group. Masticatory dysfunction was observed in 11 patients in the low PNI group and 19 patients in the high PNI group.

Reduced occlusal force was observed in 11 patients in the low PNI group and 19 patients in the high PNI group. Swallowing dysfunction was observed in five patients in the low PNI group and eight patients in the high PNI group (Table 2). Univariate analysis showed significant differences in CRP, neutrophils and tongue pressure between the two groups (p<0.01; Table 3). Multivariate analysis showed that CRP (odds ratio: 0.12, 95% confidence interval [CI]: 0.30–0.45, p<0.01) and tongue pressure (odds ratio: 3.62, 95% CI: 1.04–12.6, p<0.05) were independent risk factors for PNI (Table 3).

**Table 2.**
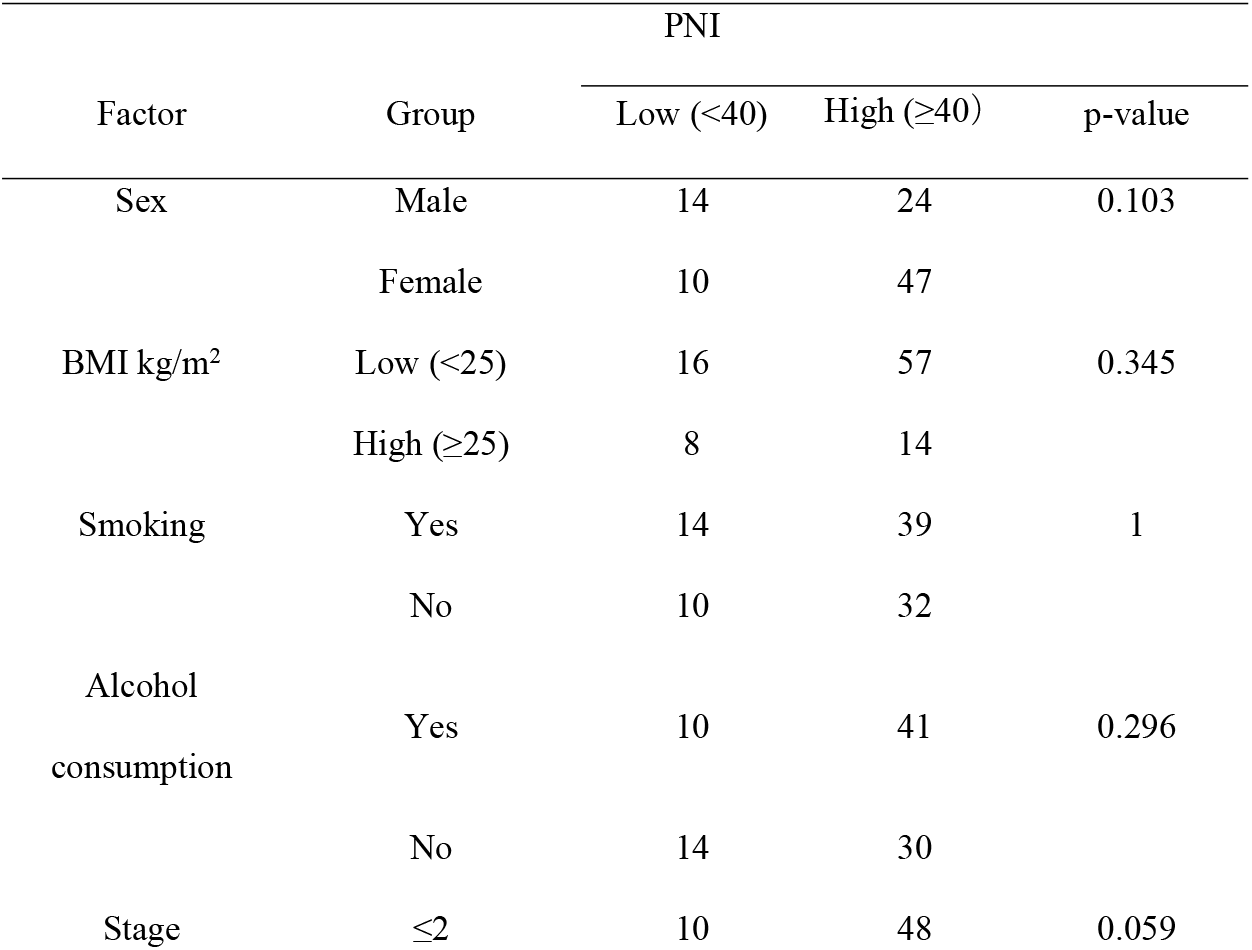
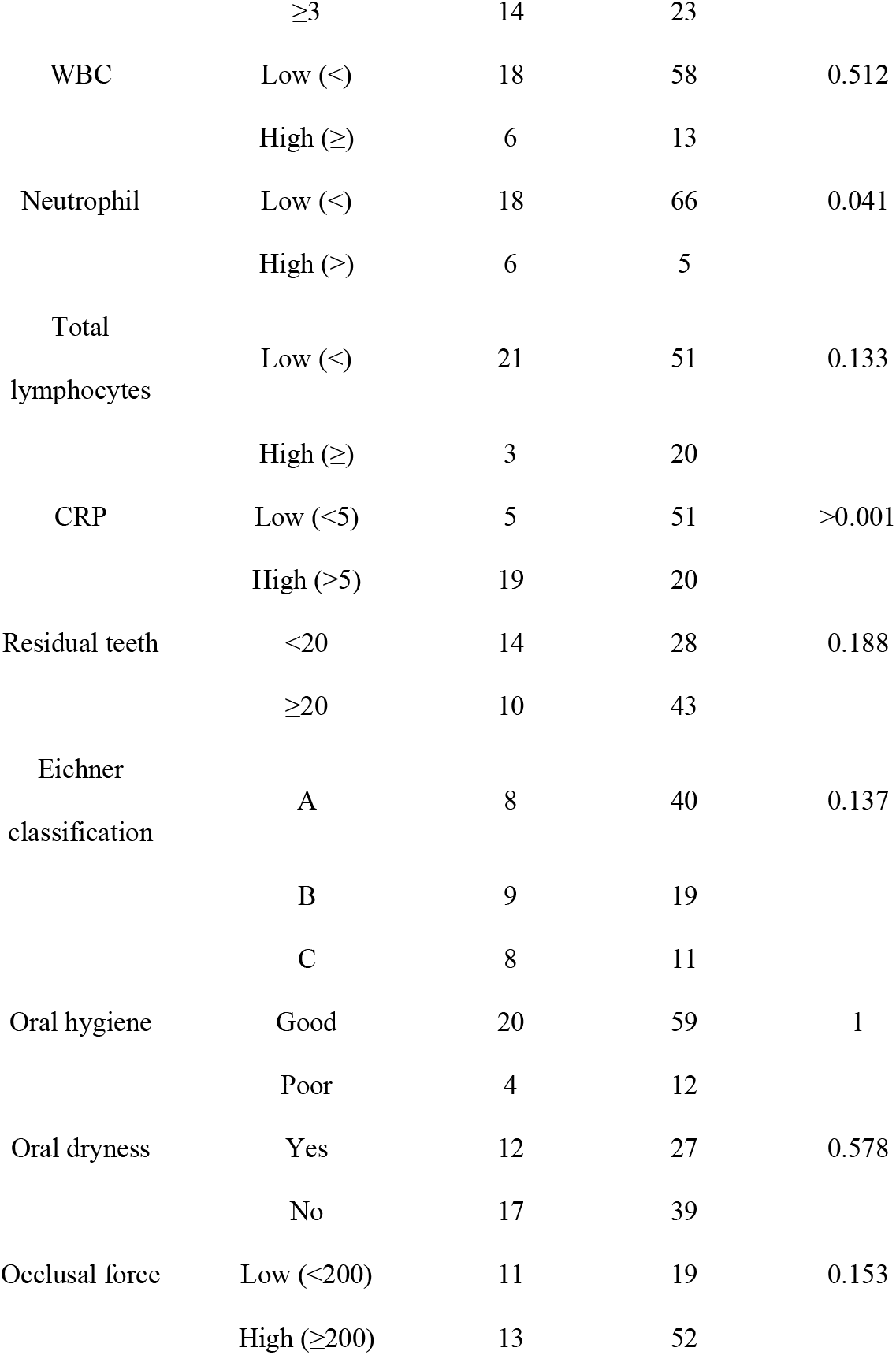
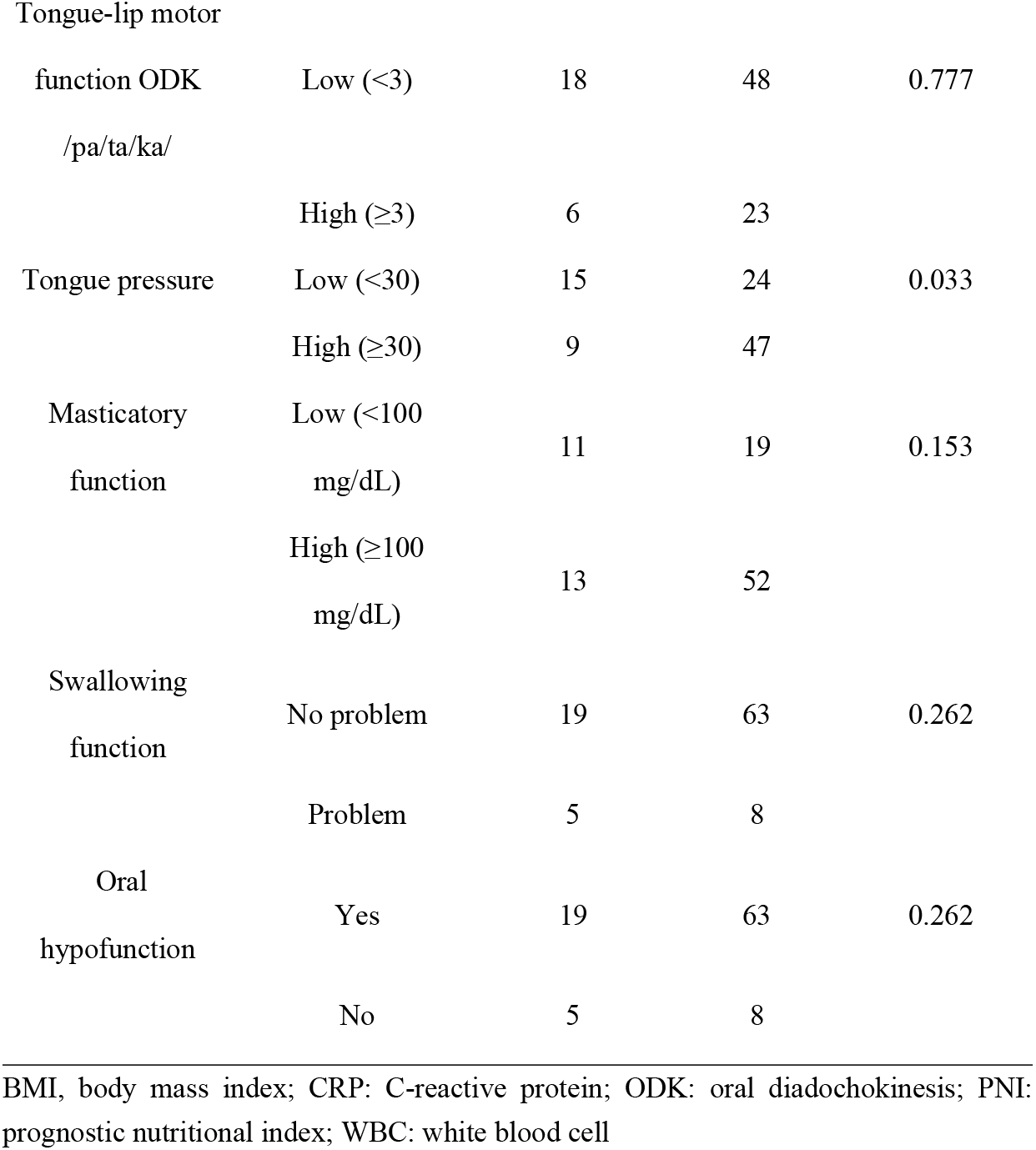
Univariate analysis.

**Table 3.**
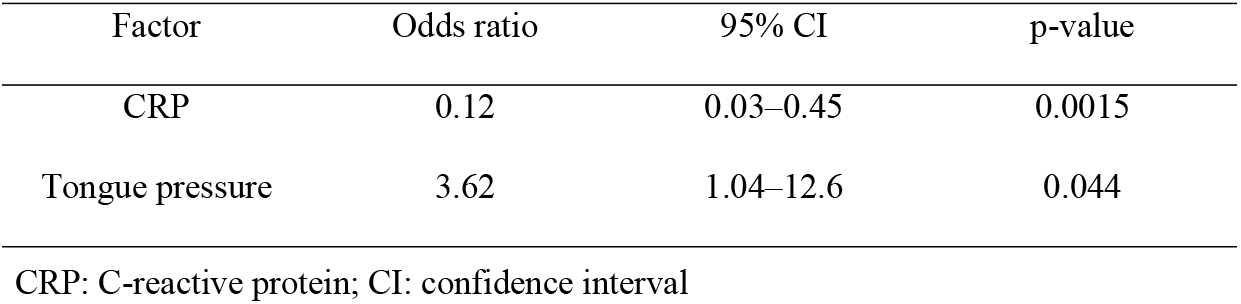
Multivariate logistic regression: Logistic regression analysis with oral hypofunction as the objective variable.

## Discussion

This cross-sectional study investigated the relationship between PNI and OHF in preoperative patients with gastric cancer. In such patients, a low PNI is an independent risk factor for poor prognosis. Consequently, patients with a low preoperative PNI must be monitored closely after surgery to avoid postoperative complications [3]. Previously, we reported that lower occlusal support levels are associated with a lower PNI [31,32]. However, the assessment of bite support range only evaluates a part of the complex oral function, such as swallowing and mastication. Regarding mastication, the risk of low nutrition and malnutrition can be prevented by devising different food forms and providing appropriate meals for patients. Therefore, in this study, we evaluated the general oral function. The results showed that high CRP and low tongue pressure were associated with a decreased PNI. Decreased tongue pressure was considered to have a possible influence on low nutrition and hidden sarcopenia. Preoperative serum CRP increases with cancer progression and is considered a prognostic indicator of cancer [1,33,34] In the microenvironment, tumour growth stimulates the production of inflammatory cytokines, such as interleukin-6 (IL-6), interleukin-8 (IL-8), and tumour necrosis factor (TNF). This is followed by inflammatory response, angiogenesis, and elevated CRP. Inflammatory cytokines, such as TNF-α, IL-1, and IL-6, also induce CRP production in the liver and cause gradual loss of critical protein components [4,5]. Thus, high CRP increases the risk of developing cancer, and the PNI is decreased when the CRP level is high. The preoperative state of nutrition in patients with gastric cancer is associated with impaired immune system function, delayed wound healing, development of complications and increased mortality [1–3]. Patients with gastric cancer are often malnourished due to stomach pain and protein leakage from tumour ulcers [2]. In addition, poor oral function can lead to loss of appetite and dysphagia, making the patient more prone to poor nutrition. Preoperative nutritional insufficiency can be caused by a variety of factors, such as OHF. Although studies have examined the relationship of nutritional status with the number of teeth loss and masticatory function [35,36], no study has related preoperative nutritional status in gastrointestinal cancer surgery to comprehensive oral functions such as swallowing, mastication, and occlusion. A diagnostic criterion consisting of seven items has been proposed by the Japanese Society of Geriatric Dentistry as an index to comprehensively evaluate the decline in oral function [16]. If the value of three of the seven items exceeds the standard value, the patient is diagnosed with decreased oral function [36]. In this study, 30 patients (40.0%) were diagnosed with decreased oral function, indicating that many patients with gastric cancer have oral function problems before surgery.

Oral functions often require compensatory movements that complement each other, it is necessary to identify the most influential factors related to oral function. Oral uncleanliness can lead to an abnormal increase in microorganisms in the oral cavity, causing pneumonia, postoperative infections, and oral infections. The higher the TCI %, the higher the number of microorganisms on the tongue. The total anaerobic bacteria count is about 7Log10 (CFU/mL) when the TCI % is about 50–60% [37]. Poor oral hygiene of the tongue diminishes the sense of taste and affects the quality of food. Specifically, poor oral hygiene may affect the intensity of taste, but it is unlikely to be directly related to nutritional status. No significant difference was found between the two groups in this study, suggesting that the effect of poor oral hygiene on PNI is small. Oral dryness alters the homeostasis of the oral environment and induces a variety of disorders. There were no significant differences in oral dryness and PNI between the groups in this study. Oral dryness may be caused by unconscious activities such as simultaneous fluid intake with food intake. Occlusal support, residual tooth count, and muscle strength affect occlusal strength [38–40]. As the number of teeth decreases, the occlusal force and masticatory capability decrease. According to many reports, oral function can be maintained if the number of remaining teeth is ≥20 and occlusal force is ≥200 N [41]. These reports suggest that a decrease in the number of teeth in patients with gastric cancer causes an alteration in diet that leads to low nutrition due to impaired gastrointestinal absorption caused by gastric cancer. In the present study, no association was found between bite force and PNI. A study reported that low occlusal force (<200 N) is associated with low body weight as well as obesity [42]. This may be because low bite strength was calculated based on a study of older people residing in institutions and the threshold was set at a lower value.

Decreased tongue and lip motor function may affect eating behaviour, nutrition, life function, and quality of life. The value of oral diadochokinesis varies depending on the condition of the oral cavity such as dentures, individual differences in ageing changes, underlying diseases, mental status, living environment, nutritional status, and presence of oral function training [43]. In the present study, no significant association was found between oral diadochokinesis and PNI. The reason may be that the frailty in patients for whom surgery had been indicated had not progressed. As surgical indications are expanded with the improvement of surgical and anaesthesia techniques, some frail patients with gastric cancer may be encountered; therefore, the effectiveness of screening needs to be additionally examined. Decreased tongue pressure due to chronic dysfunction of the lingual muscle groups decreases the pressure created between the tongue, palate, and food. The tongue performs complex mastication, swallowing, and phonation movements in coordination with the lips, mandible, pharynx, and larynx. These are important functions for maintaining life and quality of life but are difficult to quantify due to the complexity of the movements. The tongue pressure test is one of the few methods that can quantify the tongue strength. Previous studies have shown that tongue pressure is positively correlated with the food intake rate and that individuals with a maximised tongue pressure of ≥30 kPa can consume regular food, while individuals with a low tongue pressure have significantly lower food intake rate and eating patterns [14,27,44]. The results of our study showed that tongue pressure was an independent risk factor for PNI. When there is poor occlusion, it is assumed that the patient eats a soft diet and swallows by crushing the food with the tongue, jaw crest, and palate. Therefore, with appropriate tongue pressure, the compensatory mechanisms of oral function would work effectively, and the PNI would not decrease. In contrast, with low tongue pressure, the tongue cannot press against the jaw crest and palate, and compensatory movements are not possible, leading to a low PNI. Therefore, low tongue pressure is a risk factor for dysphagia and low nutrition and may also affect eating patterns, assessment and interventions such as appropriate and adequate exercise therapy, and therapeutic intervention such as improvement of oral morphology with prosthetic devices (e.g. tongue contact assisted floor), which are important for the prevention of low nutrition and muscle weakness. Dysphagia precedes the onset of obvious disability. In the absence of a causative disease, some cases of dysphagia are not recognised as diseases and do not warrant medical attention, and many symptoms, such as mild pharyngeal discomfort and swelling, occur in the older population. Therefore, the presence of hypophagia is a criterion for evaluating dysfunctional states prior to the onset of obvious disability. This reflects the patient’s perception, but there was no significant difference between the two PNI groups, which suggests that few patients eligible for surgery have significant functional decline.

Several limitations exist in this study. The present study was a single-centre study with a retrospective design, and had a small sample size. Selection bias exists because this is an observational study of surgical patients. Nutritional status is influenced by several factors, including oral findings, smoking history, psychiatric and psychological status, socioeconomic status, performance of daily activities, level of education, prevalence of processed food intake, and cultural differences in diet among countries. All of these factors were not investigated in this study. In addition, as the present study was a single-centre cross-sectional study, it was not feasible to establish a causal relationship from the findings obtained. PNI is formulated on the basis of albumin levels and lymphocyte counts. Therefore, caution should be exercised when interpreting results from patients with preoperative chemotherapy or severe inflammatory or autoimmune disease. When the diagnostic criteria for OHF are considered as a screening stage before functional impairment and malnutrition occur, it is possible to detect and respond early to the transition from OHF to malnutrition and to the flail cycle in the dental field. This early response can only be achieved through cooperation among multiple physicians and through cooperation among nurses, pharmacists, nutritionists, dental hygienists, and linguists, which will increase the contribution of dentistry to cooperation with multidisciplinary physicians and multidisciplinary professionals such as nurses, pharmacists, nutritionists, dental hygienists, speech therapists, and physical therapists.

## Data Availability

All data generated or analysed during this study are included in this published article.

## Acknowledgements

We would like to thank Editage (www.editage.com) for English language editing.

## References

1. Elahi MM, McMillan DC, McArdle CS, Angerson WJ, Sattar N. Score based on hypoalbuminemia and elevated C-reactive protein predicts survival in patients with advanced gastrointestinal cancer. Nutr Cancer. 2004;48: 171–173. doi:10.1207/s15327914nc4802_6.

2. Kushiyama S, Sakurai K, Kubo N, Tamamori Y, Nishii T, Tachimori A, et al. The preoperative geriatric nutritional risk index predicts postoperative complications in elderly patients with gastric cancer undergoing gastrectomy. In Vivo. 2018;32: 1667–1672. doi: 10.21873/invivo.11430.

3. Xishan Z, Ye Z, Feiyan M, Liang X, Shikai W. The role of prognostic nutritional index for clinical outcomes of gastric cancer after total gastrectomy. Sci Rep. 2020;10: 17373. doi: 10.1038/s41598-020-74525-8.

4. Diakos CI, Charles KA, McMillan DC, Clarke SJ. Cancer-related inflammation and treatment effectiveness. Lancet Oncol. 2014;15: e493–e503. doi: 10.1016/S1470-2045(14)70263-3.

5. Roxburgh CS, McMillan DC. Cancer and systemic inflammation: Treat the tumour and treat the host. Br J Cancer. 2014;110: 1409–1412. doi: 10.1038/bjc.2014.90.

6. Guner A, Kim HI. Biomarkers for evaluating the inflammation status in patients with cancer. J Gastric Cancer. 2019;19: 254–277. doi: 10.5230/jgc.2019.19.e29.

7. Fanetti G, Polesel J, Fratta E, Muraro E, Lupato V, Alfieri S, et al. Prognostic nutritional index predicts toxicity in head and neck cancer patients treated with definitive radiotherapy in association with chemotherapy. Nutrients. 2021;13: 127. doi: 10.3390/nu13041277.

8. Toyokawa G, Kozuma Y, Matsubara T, Haratake N, Takamori S, Akamine T, et al. Prognostic impact of controlling nutritional status score in resected lung squamous cell carcinoma. J Thorac Dis. 2017;9: 2942–2951. doi: 10.21037/jtd.2017.07.108.

9. Wu X, Jiang Y, Ge H, Diao P, Wang D, Wang Y, et al. Predictive value of prognostic nutritional index in patients with oral squamous cell carcinoma. Oral Dis. 2020;26: 903–911. doi: 10.1111/odi.13318.

10. Xue Y, Zhou X, Xue L, Zhou R, Luo J. The role of pretreatment prognostic nutritional index in esophageal cancer: A meta-analysis. J Cell Physiol. 2019;234: 19655–19662. doi: 10.1002/jcp.28565.

11. Jiang N, Deng JY, Ding XW, Ke B, Liu N, Zhang RP, et al. Prognostic nutritional index predicts postoperative complications and long-term outcomes of gastric cancer. World J Gastroenterol. 2014;20: 10537–10544. doi: 10.3748/wjg.v20.i30.10537.

12. de Marchi RJ, Hugo FN, Hilgert JB, Padilha DMP. Association between oral health status and nutritional status in south Brazilian independent-living older people. Nutrition. 2008;24: 546–553. doi: 10.1016/j.nut.2008.01.054.

13. Soini H, Muurinen S, Routasalo P, Sandelin E, Savikko N, Suominen M, et al. Oral and nutritional status-is the MNA a useful tool for dental clinics. J Nutr Health Aging. 2006;10: 495–499; discussion 500.

14. Yoshida M, Kikutani T, Tsuga K, Utanohara Y, Hayashi R, Akagawa Y. Decreased tongue pressure reflects symptom of dysphagia. Dysphagia. 2006;21: 61–65. doi: 10.1007/s00455-005-9011-6.

15. Hatanaka Y, Furuya J, Sato Y, Uchida Y, Shichita T, Kitagawa N, et al. Associations between oral hypofunction tests, age, and sex. Int J Environ Res Public Health. 2021;18: 10256. doi: 10.3390/ijerph181910256.

16. Minakuchi S, Tsuga K, Ikebe K, Ueda T, Tamura F, Nagao K, et al. Oral hypofunction in the older population: Position paper of the Japanese Society of Gerodontology in 2016. Gerodontology. 2018;35: 317–324. doi: 10.1111/ger.12347.

17. Iwasaki M, Motokawa K, Watanabe Y, Shirobe M, Ohara Y, Edahiro A, et al. Oral hypofunction and malnutrition among community-dwelling older adults: Evidence from the Otassha study. Gerodontology. 2022;39: 17–25. doi: 10.1111/ger.12580.

18. Kugimiya Y, Iwasaki M, Ohara Y, Motokawa K, Edahiro A, Shirobe M, et al. Relationship between oral hypofunction and sarcopenia in community-dwelling older adults: The Otassha study. Int J Environ Res Public Health. 2021;18: 6666. doi: 10.3390/ijerph18126666.

19. Nakamura M, Hamada T, Tanaka A, Nishi K, Kume K, Goto Y, et al. Association of oral hypofunction with frailty, sarcopenia, and mild cognitive impairment: A cross-sectional study of community-dwelling Japanese older adults. J Clin Med. 2021;10: 1626. doi: 10.3390/jcm10081626.

20. Shimazaki Y, Nonoyama T, Tsushita K, Arai H, Matsushita K, Uchibori N. Oral hypofunction and its association with frailty in community-dwelling older people. Geriatr Gerontol Int. 2020;20: 917–926. doi: 10.1111/ggi.14015.

21. Pinato DJ, North BV, Sharma R. A novel, externally validated inflammation-based prognostic algorithm in hepatocellular carcinoma: The prognostic nutritional index (PNI). Br J Cancer. 2012;106: 1439–1445. doi: 10.1038/bjc.2012.92.

22. Buzby GP, Mullen JL, Matthews DC, Hobbs CL, Rosato EF. Prognostic nutritional index in gastrointestinal surgery. Am J Surg. 1980;139: 160–167. doi: 10.1016/0002-9610(80)90246-9.

23. Onodera T, Goseki N, Kosaki G. Prognostic nutritional index in gastrointestinal surgery of malnourished cancer patients. Nihon Geka Gakkai Zasshi. 1984;85: 1001–1005. (in Japanese).

24. Shimizu T, Ueda T, Sakurai K. New method for evaluation of tongue-coating status. J Oral Rehabil. 2007;34: 442–447. doi: 10.1111/j.1365-2842.2007.01733.x.

25. Fukushima Y, Yoda T, Kokabu S, Araki R, Murata T, Kitagawa Y, et al. Evaluation of an oral moisture-checking device for screening dry mouth. Open Journal of Stomatology. 2013;3: 440–446. doi: 10.1002/cre2.145.

26. Yamada H, Nakagawa Y, Nomura Y, Yamamoto K, Suzuki M, Watanabe NY, et al. Preliminary results of moisture checker for Mucus in diagnosing dry mouth. Oral Dis. 2005;11: 405–407. doi: 10.1111/j.1601-0825.2005.01136.x.

27. Tsuga K, Yoshikawa M, Oue H, Okazaki Y, Tsuchioka H, Maruyama M, et al. Maximal voluntary tongue pressure is decreased in Japanese frail elderly persons. Gerodontology. 2012;29: e1078.#x2013;e1085-e1085. doi: 10.1111/j.1741-2358.2011.00615.x.

28. Deftereos I, Kiss N, Isenring E, Carter VM, Yeung JM. A systematic review of the effect of preoperative nutrition support on nutritional status and treatment outcomes in upper gastrointestinal cancer resection. Eur J Surg Oncol. 2020;46: 1423–1434. doi: 10.1016/j.ejso.2020.04.008.

29. Kanda Y. Investigation of the freely available easy-to-use software ‘EZR’ for medical statistics. Bone Marrow Transplant. 2013;48: 452–458. doi: 10.1038/bmt.2012.244.

30. Yang Y, Gao P, Song Y, Sun J, Chen X, Zhao J, et al. The prognostic nutritional index is a predictive indicator of prognosis and postoperative complications in gastric cancer: A meta-analysis. Eur J Surg Oncol. 2016;42: 1176–1182. doi: 10.1016/j.ejso.2016.05.029.

31. Abe A, Kurita K, Hayashi H, Ishihama T, Ueda A. Correlation between prognostic nutritional index and occlusal status in gastric cancer. Oral Dis. 2020;26: 465–472. doi: 10.1111/odi.13242.

32. Abe A, Ito Y, Hayashi H, Ishihama T, Momokita M, Taniguchi S. Correlation between geriatric nutritional risk index and oral condition in gastric cancer patients. Oral Dis. 2021. doi: 10.1111/odi.14035.

33. Erlinger TP, Platz EA, Rifai N, Helzlsouer KJ. C-reactive protein and the risk of incident colorectal cancer. JAMA. 2004;291: 585–590. doi: 10.1001/jama.291.5.585.

34. Kinoshita A, Onoda H, Imai N, Iwaku A, Oishi M, Fushiya N, et al. Comparison of the prognostic value of inflammation-based prognostic scores in patients with hepatocellular carcinoma. Br J Cancer. 2012;107: 988–993. doi: 10.1038/bjc.2012.354.

35. Hakeem FF, Bernabé E, Sabbah W. Association between oral health and frailty: A systematic review of longitudinal studies. Gerodontology. 2019;36: 205–215. doi: 10.1111/ger.12406.

36. Iwasaki M, Hirano H, Ohara Y, Motokawa K. The association of oral function with dietary intake and nutritional status among older adults: Latest evidence from epidemiological studies. Jpn Dent Sci Rev. 2021;57: 128–137. doi: 10.1016/j.jdsr.2021.07.002.

37. Iyota K, Mizutani S, Oku S, Asao M, Futatsuki T, Inoue R, et al. A cross-sectional study of age-related changes in oral function in healthy Japanese individuals. Int J Environ Res Public Health. 2020;17: 1376. doi: 10.3390/ijerph17041376.

38. Iinuma T, Arai Y, Fukumoto M, Takayama M, Abe Y, Asakura K, et al. Maximum occlusal force and physical performance in the oldest old: The Tokyo oldest old survey on total health. J Am Geriatr Soc. 2012;60: 68–76. doi: 10.1111/j.1532-5415.2011.03780.x.

39. Ikebe K, Matsuda K, Murai S, Maeda Y, Nokubi T. Validation of the Eichner index in relation to occlusal force and masticatory performance. Int J Prosthodont. 2010;23: 521–524.

40. Inomata C, Ikebe K, Kagawa R, Okubo H, Sasaki S, Okada T, et al. Significance of occlusal force for dietary fibre and vitamin intakes in independently living 70-year-old Japanese: From SONIC Study. J Dent. 2014;42: 556–564. doi: 10.1016/j.jdent.2014.02.015.

41. de Andrade FB, Lebrão ML, Santos JL, Duarte YA. Relationship between oral health and frailty in community-dwelling elderly individuals in Brazil. J Am Geriatr Soc. 2013;61: 809–814. doi: 10.1111/jgs.12221.

42. Ikebe K, Matsuda K, Morii K, Nokubi T, Ettinger RL. The relationship between oral function and body mass index among independently living older Japanese people. Int J Prosthodont. 2006;19: 539–546.

43. Umemoto G, Tsuboi Y, Kitashima A, Furuya H, Kikuta T. Impaired food transportation in Parkinson’s disease related to lingual bradykinesia. Dysphagia. 2011;26: 250–255. doi: 10.1007/s00455-010-9296-y.

44. Utanohara Y, Hayashi R, Yoshikawa M, Yoshida M, Tsuga K, Akagawa Y. Standard values of maximum tongue pressure taken using newly developed disposable tongue pressure measurement device. Dysphagia. 2008;23: 286–290. doi: 10.1007/s00455-007-9142-z.

